# Heart rate changes associated with autonomic dysreflexia in daily life of individuals with chronic spinal cord injury

**DOI:** 10.1101/2021.05.08.21256033

**Authors:** Belinda Yee, Tom E. Nightingale, Andrea L. Ramirez, Matthias Walter, Andrei V. Krassioukov

**Affiliations:** International Collaboration on Repair Discoveries (ICORD), Faculty of Medicine, University of British Columbia (UBC), Vancouver, British Columbia (BC), Canada; School of Sport, Exercise & Rehabilitation Sciences, University of Birmingham, UK; Department of Urology, University Hospital Basel, University of Basel, Basel, Switzerland; G.F. Strong Rehabilitation Centre, Vancouver, BC, Canada; Division of Physical Medicine and Rehabilitation, Faculty of Medicine, UBC, Vancouver, BC, Canada

**Author notes:** **Corresponding author**: Andrei V. Krassioukov, MD, PhD, FRCPC, ICORD, Faculty of Medicine, UBC, 818 West 10th Avenue, Vancouver, BC, V5Z 1M9, Canada. shared senior authorship (Matthias Walter, Andrei V. Krassioukov).

**Keywords:** ambulatory monitoring, autonomic dysreflexia, blood pressure, heart rate, spinal cord injury

## Abstract

**Objective:** To characterize heart rate (HR) changes during autonomic dysreflexia (AD) in daily life for individuals with chronic spinal cord injury (SCI).

**Design:** Data analysis of two prospective clinical studies and one cross-sectional study.

**Setting:** Single-center study.

**Participants:** Forty-five individuals (including 8 females) with a chronic SCI at or above the sixth thoracic spinal segment with confirmed AD, and a median age and time since injury of 43 years (interquartile range [IQR] 36 – 50) and 17 years (IQR 6 – 23) respectively, were included for analysis.

**Interventions:** Not applicable.

**Main outcome measure:** Any systolic blood pressure (SBP) increase ≥ 20mmHg from baseline from a 24-hour ambulatory surveillance period was identified and categorized as either confirmed (i.e. known AD trigger), unknown (i.e. no diary entry), and unlikely (i.e. potential physical activity driven SBP increase). SBP-associated HR changes were categorized as either unchanged, increased or decreased compared to baseline.

**Results:** A total of 797 episodes of SBP increase above AD threshold were identified and classified as confirmed (n = 250, 31.4%), unknown (n = 472, 59.2%) or unlikely (n = 75, 9.4%). Median SBP changes and median SBP-related HR changes were 37 mmHg and -8 bpm, 28 mmHg and -6 bpm, or 30 mmHg and -4 bpm for confirmed, unknown, or unlikely episodes, respectively. HR-decrease/increase ratio was 3:1 for confirmed and unknown, and 1.5:1 for unlikely episodes. HR changes resulting in brady-/tachycardia were 34.4% / 2.8% for confirmed, 39.6% / 3.4% unknown, and 26.7% / 9.3% for unlikely episodes, respectively.

**Conclusions:** Our findings suggest that the majority of confirmed AD episodes are associated with a HR decrease. Further improvements, such as more precise participant diaries combined with the use of 24-hour Holter electrocardiogram and wearable-sensors-derived measures of physical activity could provide a better, more detailed characterization of HR changes during episodes of AD.

## INTRODUCTION

There are currently 2.5 million individuals living with spinal cord injury (SCI) worldwide who face a plethora of challenges that can drastically affect their day to day quality of life^1^. Changes in certain physiological functions result in challenges such as overcoming mobility limitations and regaining control of autonomic functions such as bowel, bladder and cardiovascular regulation^2^. Cardiovascular dysfunction remains one of the leading causes of mortality and morbidity^3^ for individuals living with SCI^4,5^. Autonomic dysreflexia (AD) is a potentially life-threatening form of cardiovascular dysfunction following SCI, which can occur >40 times a day^6^. Typically this condition occurs in individuals with SCI at the 6th thoracic spinal segment (T6) or above^6^. AD is characterized by an increase in systolic blood pressure (SBP) of ≥ 20 mmHg from the individual’s baseline SBP and is elicited by noxious (e.g. soft tissue laceration or bone fracture) or innocuous (e.g. tight clothing, constipation, and full bladder) stimuli below the level of SCI^6^. Consequently, it is not uncommon for individuals with SCI to experience episodes of AD during daily bowel and bladder related care routines, with the latter being the leading cause of AD in most individuals with SCI^7^. Moreover, individuals experiencing a refractory episode of AD are at a high risk for immediate life-threatening cardio- and cerebrovascular events (e.g. stroke, brain hemorrhage, and heart attack) due to sudden spikes in SBP that can sometimes rise above 200 mmHg^5^. Although changes in SBP during AD are well defined, studies in the literature show contradicting observations of heart rate (HR) changes during AD, which suggests potential oversights in the current assessment tools for AD^8,9^. Thus, a reliable cardiovascular profile for individuals living with SCI and experiencing AD remains elusive. Our aim was to investigate the changes in HR that commonly occur during AD by analyzing not only quantitative cardiovascular parameters (e.g. SBP and HR) but also qualitative parameters (e.g. symptoms experienced and diary entries) collected from individuals with SCI at or above the T6 level during a 24-hour ambulatory surveillance period. We hope our results will contribute to building a more comprehensive cardiovascular profile for individuals suffering from AD that can guide the refinement of assessment tools and development of diagnostic tools for AD. This will ultimately help improve quality of life in this population by reducing the risks for adverse life-threatening events related to AD and complications experienced during routine bowel and bladder management protocols.

## METHODS

### Study design and ethics

This is a secondary analysis of baseline data collected for screening purposes of two prospective clinical studies (identifier NCT02298660 and NCT02676154) and one cross-sectional study, previously approved by the University of British Columbia Clinical Research Ethics Board, Vancouver Coastal Health Research Institute and Health Canada.

### Participants

The inclusion criteria for all three studies were very similar: female or male individuals at the age of 18 years or older with a chronic traumatic SCI (≥ 1 year post injury) at or above the 6^th^ thoracic spinal segment with a documented history of AD (please see supplement 1 for details). Neurological level (NLI) and severity of injury were classified in accordance with International Standards for Neurological Classification of Spinal Cord Injury (ISNCSCI)^10^.

### Outcome objective

Characterization of HR changes during 24-hour ambulatory surveillance.

### 24-hour ambulatory surveillance

All participants underwent 24-hour ambulatory blood pressure and HR monitoring (24-hour ABPM) using the Meditech Card(X)plore device (Meditech Ltd., Budapest, Hungary), a well validated tool for measuring cardiovascular profiles in individuals with SCI^11^. Both SBP and HR were programmed to be automatically recorded at 15-minute intervals during daytime (7:00 - 23:00) and at 1-h intervals at night (23:01 - 6:59). Participants also had the option to manually record their SBP and HR at any time if they were experiencing potentially AD-related signs and symptoms or were to partake in an activity that could potentially cause changes in SBP (e.g. bladder and bowl routine, physical activity).

Baseline SBP and HR values with which the 24-hour ABPM values were compared to were established as the average of three measurements taken from the participants’ nondominant arm while seated in their own wheelchairs. In addition, participants were asked to diarize any activities that could result in AD or non-AD related changes in SBP as well as potential AD signs and symptoms experienced during the 24-hour ABPM period in a participant diary.

### Data extraction

From the 24-hour ABPM records for each participant, we identified any recording with an SBP increase ≥ 20 mmHg from baseline, which is based on the clinical definition of AD as outlined by the International Standards for the Assessment of Autonomic Function after Spinal Cord Injury (ISAFSCI)^12^. We then used diarized information from the participant to categorize each episode into one of three groups: confirmed AD (known trigger was diarized, e.g. bowel routine), unknown AD (no trigger diarized), and potentially not AD (activity was diarized that could potentially cause physiological increase in SBP, e.g. physical exercise).

### Statistical analysis

Data are presented as raw values and percentages. Furthermore, data were assessed for normal distribution using the Kolmogorov-Smirnov test. Thus, results are presented as median with interquartile range. In addition, range (i.e., min - max) is presented for age and time since injury. Statistical analyses were conducted using R Statistical Software Version 3.6.0 for Mac OS.

## RESULTS

### Participants

Data from 45 participants, including 8 females, were included for analysis (Figure 1). Median age and time since injury were 43 years (36 – 50 years, range 22 – 63 years) and 17 years (6 – 23 years, range 1 – 45 years) respectively. Participant demographics and SCI characteristics are summarized in Table 1.

**Figure 1.**
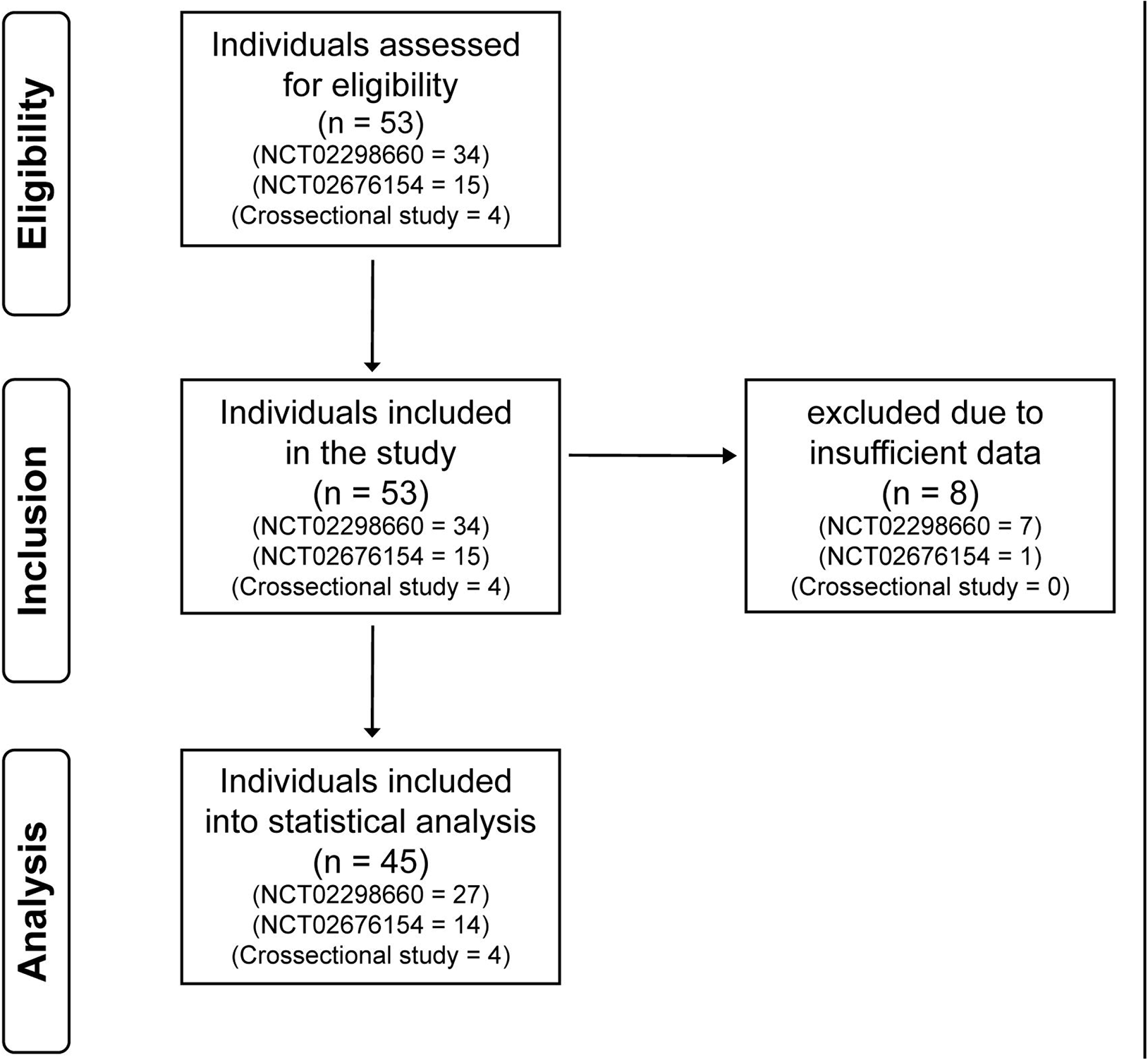
Study Flow Diagram.

**Table 1.** Demographics and injury characteristics.

| Characteristics | All (n = 45) | Cervical (n = 33) | Thoracic (n = 12) |
| --- | --- | --- | --- |
| Age [years] | 43 (36 – 50, 22 – 63) | 43 (34 – 47, 22 – 62) | 48 (40.5 – 54.25, 22 – 63) |
| Time after injury [years] | 17 (6 – 23, 1 – 45) | 16 (6 – 22, 1 – 40) | 22.5 (10 – 34.5, 1 – 45) |
| **Sex** |  |  |  |
| Female | 8 (18%) | 6 (18%) | 2 (17%) |
| Male | 37 (82%) | 27 (82%) | 10 (83%) |
| **Severity of lesion** |  |  |  |
| AIS A | 23 (51%) | 12 (36%) | 11 (92%) |
| AIS B | 16 (36%) | 15 (46%) | 1 (8%) |
| AIS C | 4 (9%) | 4 (12%) | - |
| AIS D | 2 (4%) | 2 (6%) | - |

Data are presented as median with interquartile range. In addition, range (i.e., min - max) is presented for age and time after injury. Age ( $p=0.199$ ) and time after injury ( $p=0.106$ ) were not significantly different between participants with cervical and thoracic SCI. AIS = American Spinal Injury Association Impairment Scale, n = number of participants, SCI = spinal cord injury

### Characteristics of SBP and HR during AD in daily life

Median baseline SBP across all participants was 105 mmHg (94 – 114). Median baseline SBP was significantly lower in the cervical (n = 33) compared to the upper-thoracic (n = 12) group [103 mmHg (92 – 108) vs. 114 mmHg (105 – 117), p=0.027]. Median baseline HR across all participants was 74 bpm (66 – 85). Median baseline HR was significantly lower in the cervical compared to the thoracic group [70 bpm (63 – 80) vs. 82 bpm (77 – 90), p=0.025]. Median max. SBP during AD across all participants was 172 mmHg (147 – 187). Median max. SBP during AD was not significantly different between both groups [175 mmHg (151 – 187) vs. 159 mmHg (145 – 178), p=0.397]. A total of 797 episodes where SBP increased above the AD threshold (≥ 20 mmHg) were identified and further categorized as AD trigger known (n = 250, 31.4 %), AD trigger unknown (n = 472, 59.2 %), or AD unlikely (n = 75, 9.4 %). Median change in SBP from baseline within a 24-hour period across all participants was 30 mmHg (24 – 40), while SBP-associated median HR change from baseline was -7 bpm (−18 – 1). Median changes in SBP from baseline and SBP-associated HR by category are illustrated in Figure 2 and 3, and furthermore dichotomized by NLI in Table 2, respectively. While episodes with AD trigger known (74 vs 24 %, 184 vs. 59) and unknown (73 vs. 24 %, 346 vs. 114) had both a HR decrease/increase ratio of 3:1, AD unlikely episodes were more balanced between HR decrease and increase (59 vs. 39 %, 44 vs. 29). Only 2 to 3% of all episodes in each group were associated with unchanged HR (AD trigger known: 7, AD trigger unknown: 12 and AD unlikely: 2).

**Figure 2.**
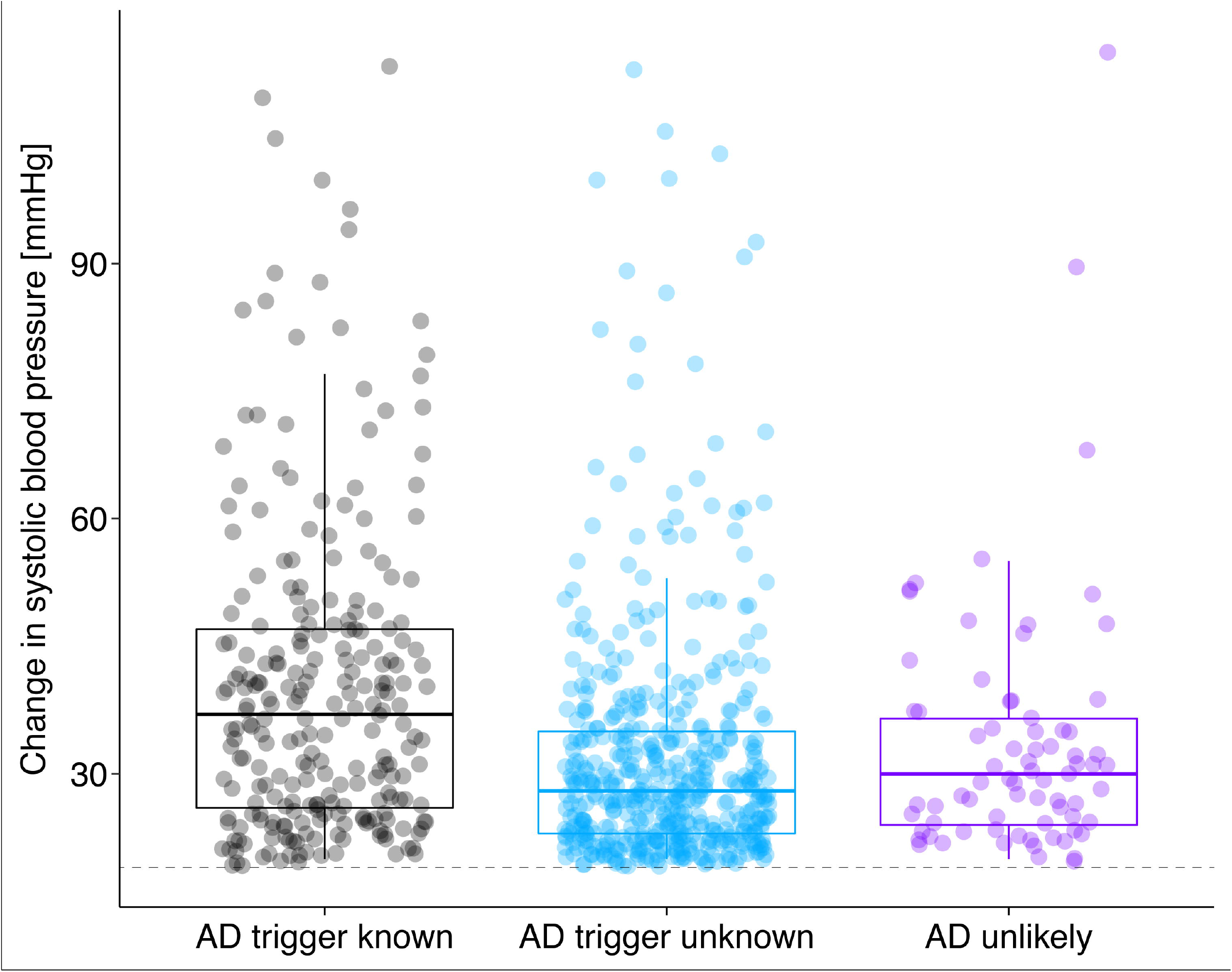
Change in systolic blood pressure from baseline during autonomic dysreflexia. Median SBP change was greatest in episodes with autonomic dysreflexia (AD) trigger known (37 mmHg, 26 – 47) compared to AD trigger unknown (28 mmHg, 23 – 35) and AD unlikely (30 mmHg, 24 – 36). Dotted line represents AD cut-off, i.e. 20mmHg SBP increase from baseline. SBP = systolic blood pressure.

**Figure 3.**
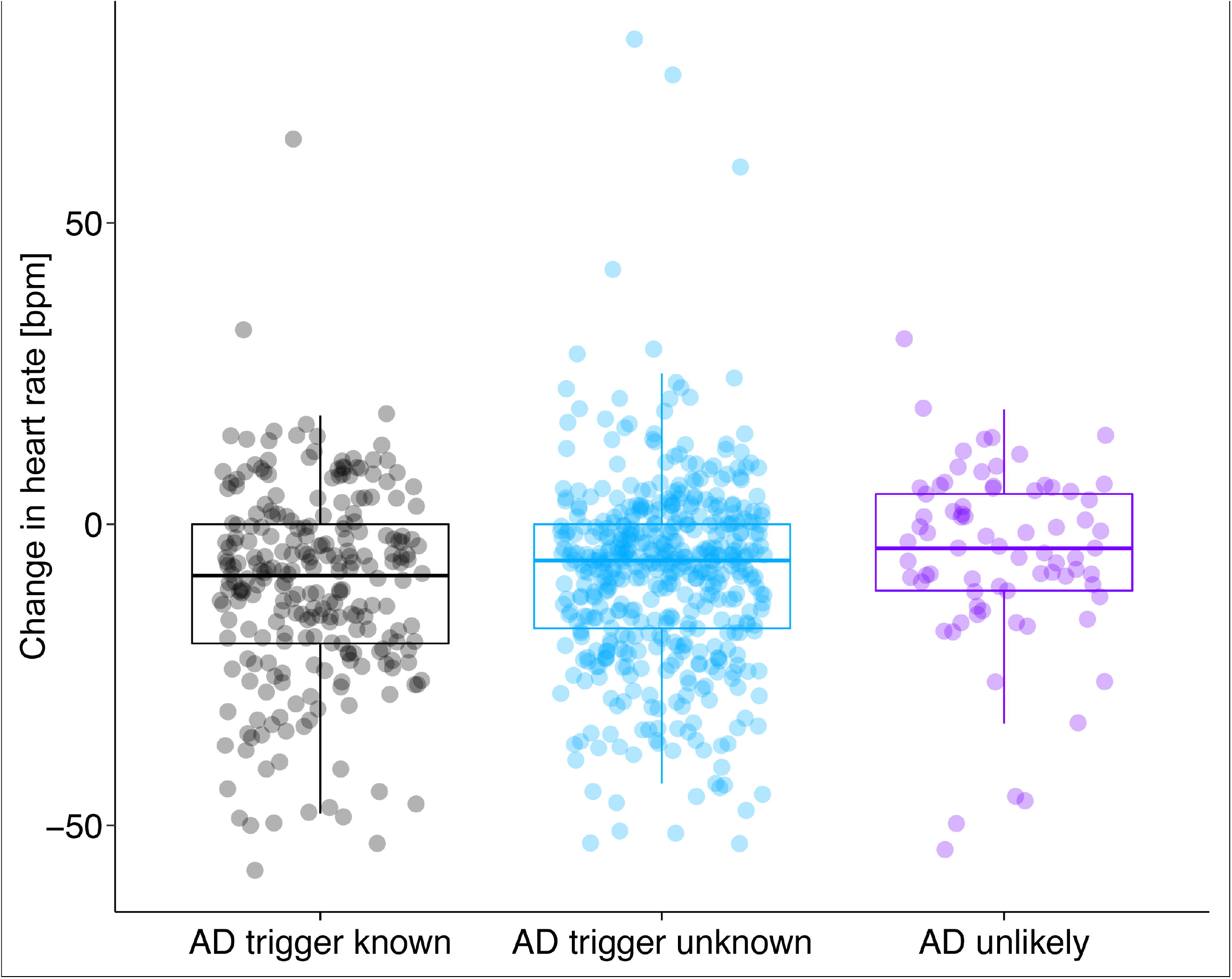
Distribution of heart rate changes associated with autonomic dysreflexia. Median heart rate (HR) change was slightly greater in episodes with autonomic dysreflexia (AD) trigger known (left, in black: -8 bpm, -20 – 0) compared to AD trigger unknown (middle, in blue: - 6 bpm, -17 – 0), and AD unlikely (right, in purple: -4 bpm, -11 – 5). BPM = beats per minute.

**Table 2.** Overall and category-specific cardiovascular changes.

| **All potential AD episodes** | **All (n<sub>e</sub> = 797)** | **Cervical (n<sub>e</sub> = 605)** | **Thoracic (n<sub>e</sub> = 192)** | **P value** |
| --- | --- | --- | --- | --- |
| SBP - Max. Δ [mmHg] | 30 (24 – 40) | 30 (24 – 41) | 27 (23 – 36) | **<0.001** |
| HR - Max. Δ [bpm] | -7 (-18 – 1) | -6 (-16 – 1) | -10 (-19 – 0) | 0.184 |
| **Known AD trigger** | **All (n<sub>e</sub> = 250)** | **Cervical (n<sub>e</sub> = 209)** | **Thoracic (n<sub>e</sub> = 42)** | **P value** |
| SBP - Max. Δ [mmHg] | 37 (26 – 47) | 38 (26 – 48) | 30 (24 – 41) | **0.006** |
| HR - Max. Δ [bpm] | -8 (-20 – 0) | -8 (-18 – 1) | -14 (-24 – -4) | 0.054 |
| **Unknown AD trigger** | **All (n<sub>e</sub> = 472)** | **Cervical (n<sub>e</sub> = 348)** | **Thoracic (n<sub>e</sub> = 123)** | **P value** |
| SBP - Max. Δ [mmHg] | 28 (23 – 35) | 28 (23 – 35) | 26 (22 – 35) | 0.091 |
| HR - Max. Δ [bpm] | -6 (-17 – 0) | -6 (-16 – 0) | -10 (-20 – -1) | 0.161 |
| **AD unlikely** | **All (n<sub>e</sub> = 75)** | **Cervical (n<sub>e</sub> = 48)** | **Thoracic (n<sub>e</sub> = 27)** | **P value** |
| SBP - Max. Δ [mmHg] | 30 (24 – 36) | 31 (26 – 37) | 28 (23 – 36) | 0.377 |
| HR - Max. Δ [bpm] | -4 (-11 – 5) | -6 (-11 – 5) | 0 (-10.5 – 5.5) | 0.284 |

Data are presented as median with interquartile range. P value indicates the comparison between those with cervical and thoracic SCI.
Δ = change, AD = autonomic dysreflexia, HR = heart rate, n = number of participants, n<sub>e</sub> = number of episodes, SBP = systolic blood pressure

### Rate of bradycardia and tachycardia at baseline and during episodes of elevated SBP

Sixteen percent of participants (7/45) had baseline HR below the threshold for bradycardia (HR < 60 bpm) and 7% (3/45) above the threshold for tachycardia (HR > 100 bpm). Number of episodes of HR changes resulting in bradycardia vs. tachycardia were 34.4% vs. 2.8% (86/250 vs. 7/250, ratio 12:1), 39.6% vs. 3.4% (187/472 vs. 16/472, ratio 12:1), and 26.7% vs. 9.3% (20/75 vs. 7/75, ratio 3:1) for AD trigger unknown, or AD unlikely episodes, respectively.

### Diarized triggers, signs and symptoms of AD

Majority of triggers associated with confirmed episodes of AD were bladder-related, with the remainder being bowel-related or related to other events (Table 3). For the episodes identified in the unlikely AD group, majority of associated activities involved *transfer to or from wheelchair* or *wheeling* (Table 4). Of the 45 participants, only 12 diarized their bodily signs and symptoms (Table 5).

**Table 3.** Frequency of diarized triggers associated with autonomic dysreflexia.

| **Trigger mechanisms of autonomic dysreflexia**<br>**with known trigger** | **Number of episodes (n<sub>e</sub> = 250)** |
| --- | --- |
| **Bladder-related** | **183 (73.2 %)** |
| Intermittent catheterization | 148 |
| Before | (50) |
| During | (68) |
| After | (30) |
| Bladder distension | 8 |
| Bladder-related discomfort/spasm | 25 |
| While draining the leg urine bag | 2 |
| **Bowel-related** | **57 (22.8 %)** |
| Before bowel stimulation | 1 |
| During bowel stimulation | 11 |
| Insertion of suppository | 2 |
| During bowel routine | 42 |
| Impacted bowel | 1 |
| **Others** | **10 (4 %)** |
| Sexual intercourse | 3 |
| Restricted clothing | 2 |
| Unspecified spasm | 5 |

**Table 4.** **Frequency of diarized activities associated with unlikely autonomic dysreflexia episodes**

| **Activities associated with unlikely autonomic dysreflexia episodes** | **Number of episodes ($n_e = 75$)** |
| --- | --- |
| Transfer (to or from bed, wheelchair, toilet, car etc.) | 46 (61.3 %) |
| Position change (lie down or sit up) | 7 (9.3 %) |
| Workout / stretching | 2 (2.7 %) |
| Wheeling | 17 (22.7 %) |
| Transferring laundry | 1 (1.3 %) |
| Getting dressed | 1 (1.3 %) |
| Anxiety | 1 (1.3 %) |

**Table 5.** **Frequency of diarized autonomic dysreflexia specific signs and symptoms and non-specific bodily symptoms** Twenty-seven percent of the participants (12/45) diarized bodily signs and symptoms experienced during the 24-hour surveillance period. A total of six participants recorded specific signs and symptoms associated with autonomic dysreflexia (AD), such as “goosebumps” (piloerection), headaches, “profound sweating” (hyperhidrosis). *Two participants recorded experiencing “AD symptoms” or “high blood pressure” but did not specify what they were (unspecified). Four participants reported only non-specific bodily symptoms that are not generally attributable to AD. Of the six participants who recorded AD specific symptoms, three of them also recorded non-specific bodily symptoms.

| **Frequency of diarized autonomic dysreflexia specific signs and symptoms** | **AD trigger known** | **AD trigger unknown** | **AD unlikely** |
| --- | --- | --- | --- |
| Piloerection | 9 | 0 | 0 |
| Headache | 4 | 1 | 1 |
| Profound sweating (hyperhidrosis) | 10 | 1 | 0 |
| Unspecified* | 2 | 5 | 0 |
| **Frequency of diarized non-specific bodily symptoms** |  |  |  |
| Stress / fatigue | 3 | 0 | 0 |
| Pain | 1 | 1 | 0 |
| Muscle spasm | 1 | 2 | 0 |
| Nausea | 1 | 0 | 0 |
| Bladder Pressure | 4 | 1 | 0 |

## DISCUSSION

The main findings from our data analysis showed that most confirmed (74%) and unknown (73%) AD episodes were associated with a HR decrease while unlikely AD episodes were more balanced with respect to HR decreases (59%) and increases (39%). Moreover, the overall rate of bradycardia associated with identified episodes of elevated SBP was much higher than that of tachycardia regardless of whether an AD trigger was identified. Confirmed episodes of AD were most frequently associated with bladder-related events.

The high rate of bradycardia and HR decreases associated with confirmed AD cases in our analysis is in line with the previous understanding of the parasympathetically mediated HR changes that are believed to occur during AD, which suggest bradycardia as a pathognomonic feature of AD^13^. In SCI, the innervation of the heart (T1-T5) may be compromised, but parasympathetic innervation of the heart by the vagus nerve remains intact as it is located external to the spinal system and more or less functioning independently from it^14^. Thus, bradycardia may be a compensatory response by the parasympathetic system to counteract the significantly elevated SBP in AD^3,14^. Although we applied the clinical definition of bradycardia in our statistical analysis, it may not be applicable as it does not take into account the new baseline cardiovascular changes that are established in individuals with chronic SCI^15^. The level of neurological lesion associated with chronic SCI may have an impact on baseline resting HR as one meta-analysis depicted resting HR to be significantly lower in individuals with chronic SCI originating at the cervical level compared to those below T6^15^. As our cohort consisted of mostly individuals with lesions of cervical origin (n = 33), this may appear to account for the high rates of bradycardia seen. Rather than applying classic clinical definitions or setting arbitrary thresholds to describe HR changes, alternate methods to describe cardiovascular parameters may more appropriately reflect the physiological baseline changes associated with chronic SCI. Analyzing the change in HR from baseline may be a more appropriate method as some of the participants in our study had baseline HR values in the bradycardic and tachycardic range.

Contrary to our results, a few studies have indicated tachycardia as a predominant feature of AD with bradycardia occurring much less frequently. Kewalramani’s combined retrospective and prospective review of AD episodes in patients with traumatic myelopathy showed bradycardia in 10 % of participants and tachycardia in 38 %, which was the opposite of our findings ^9^. Solinsky et al.’s analysis of a prospectively acquired AD dataset showed a significantly higher prevalence of tachycardia during AD as 68 % of AD episodes were associated with tachycardia while 0.3 % were associated with bradycardia^8^. Lindan et al.’s study found bradycardia and tachycardia to be equally prevalent during AD^16^. Karlsson speculated the presence of tachycardia in AD to be due to spinal signals travelling back up the tract to instigate the sympathetic activation of the heart, causing an increase in heart rate^13^. He suggested a marked difference seen in the HR responses of individuals with cervical vs. thoracic lesions would support this explanation^13^. In our analysis, mean HR change between those with thoracic and cervical SCI were not significantly different (Table 2) which would not be in line with this proposed theory.

Potential explanations for the relative tachycardia seen predominantly in other studies could include misidentification of AD due to lack of contextual information, confounding variables such as systemic comorbidities, medications affecting the cardiovascular system, and lack of true baseline measurements etc. As the core strength of our study lies within its methodology, these factors were accounted for through the design as well as the exclusion and inclusion criteria for the clinical trials included in our analysis. While past studies have relied on SBP thresholds (e.g. 150 mmHg) that differ from the clinical definition of AD or AD symptomology to identify episodes of AD, our study used ISAFSCI’s definition of AD for identification^8,9,16^. This means our methodology was designed to be more robust in identification of low-grade AD and asymptomatic AD episodes, as well as reduce the likelihood of misidentifying elevated SBP readings due to essential hypertension as AD. Furthermore, our study utilized 24-hour ABPM data in conjunction with diary entries to provide detailed context, which optimized validity of AD identification by capturing commonly experienced habitual AD triggers over a prolonged period in each participant’s everyday routine rather than in one point in time. This contrasts with the Kewalramani, Solinsky, and Lindan studies, which analyzed AD episodes in an inpatient setting where the context surrounding their admission and the treatments or procedures they underwent remained largely unknown, unless it was related to AD. Detailed context is important as certain provocations such as laryngoscopy and tracheal intubation have been shown to elicit arrhythmias and tachycardia^17^. In addition, our study determined baseline vitals prior to the initiation of any treatment. Previous studies in the literature analyzed inpatient data and may have used baseline vitals for comparison that were determined after the initiation of treatments or not used baseline values at all^8,9,16^.

The analysis of participants diaries allowed for the identification of activities that were true triggers of AD and for confounding activities that mimicked the SBP elevation in AD but are considered false triggers. The most common triggers of confirmed AD episodes noted in participant diary entries (Table 3) were associated with bladder-related events, which is in line with literature reports as the most common trigger of AD^7,16^. Examples of diarized confounding activities that are not considered triggers of AD (Table 4) but concurrently correlated with episodes of elevated SBP (thus categorized into the unlikely AD group) included anxiety and physical activity during exercise, wheeling, or transfers to/from wheelchair. The misidentification of AD was discussed recently in 2 case studies where participants undergoing urodynamic studies had elevated SBP thought to be associated with AD^18^. Further review revealed that the elevation in SBP was likely due to anxiety as SBP decrease was noted after participants were informed of the cancellation or completion of the study but prior to the initiation of AD therapeutic interventions^18^. Such confusion surrounding AD identification can result in a misunderstanding of its accompanying cardiovascular parameters. These events can be concurrently monitored through the self-report diary or captured by utilizing a combination of wearable devices to optimize the identification of AD episodes and cardiovascular parameters during free-living monitoring. For example, a 24-hr AMPM device to capture BP, 24-hour Holter electrocardiogram to capture HR and sensors/accelerometers to capture physical activity to rule out a non-AD mediated increase in SBP. In the future, such approaches may help to optimize the provision of healthcare for SCI patients by increasing awareness regarding the importance of AD recognition, since a significant lack of knowledge in this domain has been exhibited in medical staff working at emergency departments^19^. Even more alarming, current literature indicates that up to 41 % of individuals with chronic SCI have never heard of AD despite 22 % reporting signs of unrecognized AD^20^.

### Study limitations

One limitation of our study was that the majority of episodes with elevated SBP ≥ 20 mmHg were recorded without associated triggers, which could, at least in part, be seen as asymptomatic AD, i.e. silent AD^21^. However, it could also partially be attributed to the variability in the quality of participant diary records, or the lack thereof. Only 16% of our participants reported experiencing AD specific symptoms and it is unknown whether those who did not record AD symptoms truly experienced no symptoms at all or failed to record them for other reasons (e.g., perceived burden of providing a detailed recall of activities over a 24-hr period). This happened despite participants undergoing an in depth debriefing with a research coordinator the day after the 24-hour surveillance period to clarify diary records and activities. Although silent AD is relatively common in the literature, occurring in more than 50% of individuals who experience AD^21,22^, educational sessions about AD sign and symptom recognition for participants prior to the 24-hour ABPM period could improve the quality of diary reports. We also did not measure physical activity to rule out non-AD mediated increases in SBP. Future studies can consider use of wearable sensors/devices to monitor energy expenditure^23^. Expanding the 24-hour ABPM and diary data collection period to multiple days rather than just a single 24-hour period would allow for a more robust analysis of cardiovascular parameters during AD in daily life and provide more insight on silent AD. However, participants compliance could decline the longer the surveillance period lasts. Another limitation was that the ratio of cervical to thoracic level lesions (3:1) as well as between motor-complete to motor-incomplete injury (>6:1) in our participants. Considering this, future studies should aim to achieve a balanced ratio of the level and severity of injury.

## CONCLUSION

Our results suggest that most confirmed AD episodes were associated with a HR decrease. However, the number of unknown episodes, which could potentially be silent AD, was high. Nonetheless, the use of 24-hour ABPM in conjunction with diary records of daily activities and symptoms provides a promising methodology for the valid identification and differentiation of confirmed AD episodes from unlikely AD episodes, which is essential for developing a more accurate cardiovascular profile for individuals with chronic SCI experiencing AD.

Developing a more accurate and comprehensive cardiovascular profile for this population can ultimately help improve quality of life by reducing the risks for adverse life-threatening cardiovascular events and complications related to AD experienced during routine daily activities, such as initiation of bowel and bladder management protocols. Further improvements, such as a more precise documentation of participant diaries and the combined use of 24-hour Holter electrocardiogram and wearable-sensors-derived measures of physical activity could increase the validity of HR changes during AD.

## Supporting information

please see supplement for details

## Data Availability

The data are available from the corresponding author on reasonable request.

## Author contributions

Andrei V. Krassioukov and Matthias Walter had full access to all the data in the study and take responsibility for the integrity of the data and the accuracy of the data analysis.

## Study concept and design

Belinda Yee, Thomas E. Nightingale, Andrea L. Ramirez, Andrei V. Krassioukov, Matthias Walter

## Acquisition of data

Belinda Yee, Thomas E. Nightingale, Andrea L. Ramirez, Andrei V. Krassioukov, Matthias Walter

## Analysis and interpretation of data

Belinda Yee, Thomas E. Nightingale, Andrea L. Ramirez, Andrei V. Krassioukov, Matthias Walter

## Drafting of the manuscript

Belinda Yee

## Critical revision of the manuscript for important intellectual content

Thomas E. Nightingale, Andrea L. Ramirez, Andrei V. Krassioukov, Matthias Walter

## Statistical analysis

Belinda Yee and Matthias Walter

## Supervision

Andrei V. Krassioukov and Matthias Walter

## Obtaining funding

Belinda Yee is a University of British Columbia - Faculty of Medicine Summer Student Research Program (FoM SSRP) award recipient. Tom Nightingale (grant number 17767) and Matthias Walter (grant number 17110) were recipients of Michael Smith Foundation for Health Research Trainee Awards in conjunction with the International Collaboration on Repair Discoveries and Rick Hansen Foundation, respectively. Dr. Krassioukov is supported by Endowed Chair, Department of Medicine, Univerty of British Columbia. NCT02298660 (Praxis Spinal Cord Institute, i.e., formerly Rick Hansen Institute, grant number G2013-09 and Allergan Inc.), NCT02676154 (Pfizer Canada investigator-initiated research, grant number WI207218) and cross-sectional study (Praxis Spinal Cord Institute, i.e., formerly Rick Hansen Institute, grant number G2015-31).

