## Supplementary material for "Heart rate changes associated with autonomic dysreflexia in daily life of individuals with chronic spinal cord injury": please see supplement for details

**Study location:**

**NCT02298660 (https://clinicaltrials.gov/ct2/show/study/NCT02298660)**

Inclusion criteria:

- Inpatients or outpatients with SCI (AIS A-D)
- Male and female
- Age between 18 - 65
- Chronic, traumatic SCI (> 1 year post injury)
- Affected by urinary incontinence
- We are expecting individuals with the following levels of injury:
- Individuals with spinal segment thoracic (T) 6 and above (with history of episodes of AD) Presence of AD will be determined using a validated AD questionnaire
- Good command and comprehension of English
- Capable of giving informed consent

Exclusion criteria:

- Age older than 66 years
- Documented traumatic brain injury
- Acute co-morbidities
- Other diseases of the neural system
- Previous genitourinary disease or operation,
- Current urinary tract infection
- Multiple injury levels
- Previous history of systemic illness, such as cardiovascular diseases (as hypertension and cardiac infarction), cerebrovascular accident, diabetes, etc.
- Poor command of English language
- Pregnancy

**NCT02676154 (https://clinicaltrials.gov/ct2/show/NCT02676154)**

Inclusion criteria:

- Male or female, 18 - 60 years of age
- Chronic traumatic SCI at or above T6 spinal segment and >1 year post injury
- Documented presence of AD and NDO during UDS
- Hand function sufficient to perform CIC or a committed caregiver to provide CIC for management of urinary bladder drainage
- Patients must have documented two weeks of bladder and bowel history prior to their baseline visit
- Willing and able to comply with all clinic visits and study-related procedures
- Able to understand and complete study-related questionnaires (must be able to understand and speak English or have access to an appropriate interpreter as judged by the investigator)
- Women of childbearing potential must not be intended to become pregnant, currently pregnant, or lactating. The following conditions apply:
    - Women of childbearing potential must have a confirmed negative pregnancy test prior to the baseline visit. During the trial, all women of childbearing potential will undergo urine pregnancy tests at their monthly clinic visits as outlined in the schedule of events
    - Women of childbearign potential must agree to use adequate contraception during the period of the trial and for at least 28 days after completion of treatment. Effective contraception includes abstinence
- Sexually active males with female partners of childbearing potential must agree to use effective contraception during the period of the trial and for at least 28 days after completion of treatment
- Must Provide Informed Consent

Exclusion criteria:

- Presence of severe acute medical issue that in the investigator's judgement would adversely affect the patient's participation in the study
- A hypersensitivity to tolterodine (available as Detrol, Detrol LA), soya, peanuts, or lactose
- Recent treatment with intravesical OnabotulinumtoxinA (within 9 months of the baseline visit)
- Recent treatment with other anticholinergics medications (within 3 weeks of the baseline visit)
- Use of any medication or treatment that in the opinion of the investigator indicates that it is not in the best interest of the patient to participate in this study
- Patient is a member of the investigational team or his /her immediate family

**Cross-sectional, exploratory study investigating the impact of blood pressure instability on cerebrovascular health in individuals with SCI compared to able-bodied control participants.**

Recruitment of age and sex-matched able-bodied control participants is still ongoing.

Inclusion criteria:

- Male or female
- 19 - 65 years of age
- Chronic traumatic motor complete SCI (AIS A or B) between C4-T6 spinal segment and >1 year post injury
- Documented presence of AD
- Participants must be competent enough to give consent
- Willing and able to comply with all clinic visits and study-related procedures
- Able to understand and complete study-related questionnaires

Exclusion criteria:

- History of:
    a. Untreated cardiovascular disease
    b. Type 1 diabetes
    c. Untreated Type 2 diabetes
    d. Obesity
    e. Severe brain injury
    f. Depression (confounder of cognition)
